## Supplemental Tables for "Investigating the role of the relaxin-3/RXFP3 system in neuropsychiatric disorders and metabolic phenotypes: a candidate gene approach"

**Supplementary Table 1**: Full description and field codes used to derive phenotype caseness definitions for depression and atypical depression.

| **Depression Phenotypes** | **Caseness description** | **Relevant data field codes** |
| --- | --- | --- |
| Broad depression | Answered “yes” to the question “Have you ever seen a GP for nerves, anxiety, tension or depression”  **OR**  Answered “yes” to the question “Have you ever seen a GP/psychiatrist for nerves, anxiety, tension or depression”  **OR**  Have a primary or secondary diagnosis of one of the following ICD-10 codes for mood disorders in hospital episode data from UK bodies:   - F32: depressive episode - F33: recurrent depressive disorder - F34: persistent mood disorders - F38: other mood disorders - F39: unspecified mood disorder | 2090, 2100  41202, 41204 |
| ICD10-coded depression | Have a primary or secondary diagnosis of one of the following ICD-10 codes for mood disorders in hospital episode data from UK bodies:   - F32: depressive episode - F33: recurrent depressive disorder - F34: persistent mood disorders - F38: other mood disorders - F39: unspecified mood disorder | 41202, 41204 |
| Lifetime depression | Indication of major depression status from derived data field generated by DJ Smith et al.^1^, based on touchscreen responses for help-seeking behaviour and extended presence of low mood or anhedonia | 20126 |
| CIDI depression | Met all the following criteria:   1. Answered “yes” to having experienced anhedonia or low mood for two weeks or more in a row 2. Answered “yes” to having 4 or more of the following 8 symptoms, based on the CIDI assessment:    a. Anhedonia    b. Low mood    c. Trouble with concentration    d. Ideation of death    e. More tired or low on energy than usual    f. Feelings of worthlessness    g. Trouble falling asleep, sleeping too much, or waking too early    h. Gaining or losing weight 3. Answered “almost every day” or “every day” [coding 3] to a question about the frequency of depressed days during their worst episode of depression. 4. Answered either “about half of the day”, “most of the day”, or “all day long” to the question “How much of the day did these feelings usually last?" during their worst episode of depression. 5. Answered “somewhat” [coding 2] or “a lot” [coding 3] to the following prompt: "Think about your roles at the time of this episode, including study/employment, childcare and housework, leisure pursuits. How much did these problems interfere with your life or activities?", during their worst episode of depression | 20446, 20441  20435, 20437, 20449, 20450, 20533, 20534, 20535, 20536  20439, 20436, 20440 |
| PHQ-9 definition depression | Questions from the PHQ-9 screening tool were administered in an online mental health follow up questionnaire.  Cases met all the following criteria:   1. Answered “more than half the days” or “nearly every day” when asked how often they had been bothered by either anhedonia or low mood. 2. Answered “more than half the days” or “nearly every day” for 5 or more of the following symptoms, when asked how often they had been bothered by said symptom:    a. Lack of interest or pleasure in doing things    b. Depression    c. Trouble concentrating on things    d. Thoughts of suicide or self-harm    e. Feelings of tiredness or low energy    f. Feelings of inadequacy    g. Trouble falling asleep, sleeping too much, or waking too early    h. Poor appetite or overeating    i. Changes in speed/amount of moving or speaking | 20514, 20510  20507, 20508, 20517, 20518, 20519, 20511, 20513 |
| PHQ-9 cutoff depression | Questions from the PHQ-9 screening tool were administered in an online mental health follow up questionnaire.  The presence of each of the following symptoms was rated on a scale of 1-4, analogous to the PHQ-9 ratings of 0-3. 1 corresponds to not at all, 2 corresponds to several days, 3 corresponds to more than half the days, and 4 corresponds to nearly every day:   1. Recent feelings of depression 2. Recent lack of interest or pleasure in doing things 3. Recent feelings of tiredness or low energy 4. Recent trouble concentrating on things 5. Recent thoughts of suicide or self-harm 6. Recent poor appetite or overeating 7. Trouble falling or staying asleep, or sleeping too much 8. Recent feelings of inadequacy 9. Recent changes in speed/amount of moving or speaking   Cases had a score of ≥19, corresponding to the PHQ-9 cutoff score of ≥10. | 20514, 20510  20507, 20508, 20517, 20518, 20519, 20511, 20513 |
| CIDI atypical depression | Met case status on the CIDI depression phenotype definition  **AND**  Answered “yes” to sleeping too much during worst period of depression or answered “gained weight” when asked about weight change during worst episode of depression. | 20446, 20441, 20435, 20437, 20449, 20450, 20533, 20534, 20535, 20536, 20439, 20436, 20440  20536, 20534 |
| PHQ-9 definition atypical depression | Met case status on the PHQ-9 definition phenotype of depression  **AND**  Answered “yes” to sleeping too much during worst period of depression or answered “gained weight” when asked about weight change during worst episode of depression. | 20514, 20510, 20507, 20508, 20517, 20518, 20519, 20511, 20513  20536, 20534 |
| PHQ-9 cutoff atypical depression | Met case status on the PHQ-9 cutoff phenotype of depression  **AND**  Answered “yes” to sleeping too much during worst period of depression or answered “gained weight” when asked about weight change during worst episode of depression. | 20514, 20510, 20507, 20508, 20517, 20518, 20519, 20511, 20513  20536, 20534 |

**Supplementary Table 2**: Full description and field codes used to derive phenotype caseness definitions for anxiety.

| **Anxiety Phenotypes** | **Description** | **Relevant field codes** |
| --- | --- | --- |
| ICD10-coded anxiety | Have a primary or secondary diagnosis of one of the seven following ICD-10 codes for anxiety disorders in hospital episode data from UK bodies:   - F40: phobic anxiety disorders - F41: other anxiety disorder - F42: obsessive-compulsive disorder - F43: reaction to severe stress, and adjustment disorders - F44: dissociative disorders - F45: somatoform disorders - F48: other neurotic disorders | 41202, 41204 |
| Lifetime disorder anxiety | Met all the following criteria:   1. Answered yes to either of the following questions: "People differ a lot in how much they worry about things. Did you ever have a time when you worried a lot more than most people would in your situation?" **or** "Please think of the period in your life when you have felt worried, tense, anxious, or more worried than most people would in your situation. This could be in the past, or it could be continuing now. During that period, was your worry stronger than in other people?" 2. Answered yes to the question "Have you ever had a period lasting one month or longer when most of the time you felt worried, tense, or anxious?" 3. Had this frequent and persistent worrying for 6 months or longer 4. Answered yes to the question “Did you worry most days?" during their worst period of anxiety 5. Answered yes to either of the following questions: " Did you ever have different worries on your mind at the same time?" **or** "Did you usually worry about one particular thing, such as your job security or the failing health of a loved one, or more than one thing?" during their worst period of anxiety 6. Answered yes to the question “Did you find it difficult to stop worrying?" **or** answered “often” to the question “How often was your worry so strong that you couldn't put it out of your mind no matter how hard you tried?" **or** answered “often” to the question “How often did you find it difficult to control your worry?" 7. Answered “a lot” to the question "Think about your roles at the time of this episode, including study / employment, childcare and housework, leisure pursuits. How much did these problems interfere with your life or activities?" 8. Suffered 3 or more somatic symptoms of anxiety during their worst period of anxiety (including feelings of restlessness; feeling easily tired; having trouble falling or staying asleep; feeling keyed up or on edge; increased irritability; experiencing tense, sore or aching muscles; having difficulty concentrating)   **OR**  Self-reported receiving a professional diagnosis of one or more of the following disorders:   - Social anxiety or social phobia - Any other phobia (eg disabling fear of heights or spiders) - Panic attacks - Obsessive compulsive disorder (OCD) - Anxiety, nerves or generalized anxiety disorder - agoraphobia | 20425, 20542, 20426, 20429, 20427, 20423, 20422, 20417, 20419  20421, 20420, 20538, 20540, 20543, 20541, 20539, 20537  20544 |
| GAD-7 cutoff anxiety | Questions from the GAD-7 screening tool were administered in an online mental health follow up questionnaire. The presence of each of the following symptoms was rated on a scale of 1-4, analogous to GAD-7 ratings of 1-3. 1 corresponds to not at all, 2 corresponds to several days, 3 corresponds to more than half the days, and 4 corresponds to nearly every day:   1. Recent easy annoyance or irritability 2. Recent feelings of foreboding 3. Recent feelings or nervousness or anxiety 4. Recent inability to stop or control worrying 5. Recent restlessness 6. Recent trouble relaxing 7. Recent worrying too much about different things   Cases had a score of ≥17, corresponding to the GAD-7 cutoff score of ≥10. | 20505, 20512, 20506, 20509, 20516, 20515, 20520 |

**Supplementary Table 3**: Full description and field codes used to derive phenotype caseness definitions for metabolic syndrome and the sub-outcomes that comprise the disorder.

| **Metabolic Phenotypes** | **Description** | **Relevant field codes** |
| --- | --- | --- |
| High waist circumference | Waist circumference of ⩾102 cm for males or ⩾88 cm for females | 48 |
| Hypertension | Systolic blood pressure ⩾130 mmHg or diastolic blood pressure ⩾85 mmHg  **OR**  On one of the following antihypertensive drug treatments, based on participant self-reported medication data:   - Doxazosin - Clonidine - Moxonidine - Indoramin - Methyldopa - Prazosin - Minoxidil - Hydralazine - Bendroflumethiazide - Atenolol - Bisoprolol - Furosemide - Propranolol - Indapamide - Spironolactone - Metoprolol - Sotalol - Chlorthalidone \|Atenolol - Amiloride \|Furosemide - Nebivolol - Bumetanide - Timolol - Carvedilol - Bisoprolol \|Hydrochlorothiazide - Amiloride \|Hydrochlorothiazide - Amiloride - Celiprolol - Bendroflumethiazide \|Potassium - Atenolol \|Bendroflumethiazide - Eplerenone - Labetalol - Triamterene \|Hydrochlorothiazide - Bendroflumethiazide \|Propranolol - Hydrochlorothiazide - Chlorthalidone - Sotalol \|Hydrochlorothiazide - Carteolol - Betaxolol - Amiloride \|Cyclopenthiazide - Atenolol \|Nifedipine - Metoprolol \|Chlorthalidone - Acebutolol - Oxprenolol - Atenolol \|Amiloride \|Hydrochlorothiazide - Nadolol - Torasemide - Metolazone - Xipamide - Pindolol - Cyclopenthiazide - Ramipril - Amlodipine - Lisinopril - Perindopril - Candesartan - Losartan - Felodipine - Irbesartan - Enalapril - Valsartan - Lercanidipine - Nifedipine - Diltiazem - Verapamil - Telmisartan - Olmesartan - Lacidipine - Losartan \|Hydrochlorothiazide - Trandolapril - Eprosartan - Fosinopril - Captopril - Quinapril - Lisinopril \|Hydrochlorothiazide - Hydrochlorothiazide \|Irbesartan - Enalapril \|Hydrochlorothiazide - Perindopril \|Indapamide - Valsartan \|Hydrochlorothiazide - Diltiazem \|Hydrochlorothiazide - Nicardipine - Telmisartan \|Hydrochlorothiazide - Ramipril \|Felodipine - Imidapril - Cilazapril - Hydrochlorothiazide \|Captopril | 4079, 4080, 20003 |
| Hypertriglyceridaemia | Triglycerides level ⩾150 mg/dL  **OR**  On one of the following drug treatments for elevated triglycerides, based on participant self-reported medication data:   - Fenofibrate - Bezafibrate - Ciprofibrate - Gemfibrozil - Ezetimibe - Fish Oil - Omega-3-Acid Ethyl Esters - Simvastatin - Atorvastatin - Rosuvastatin - Pravastatin - Fluvastatin | 30870, 20003 |
| Low HDL cholesterol  (Dyslipidaemia) | HDL cholesterol levels < 40 mg/dL for men or < 50 mg/dL for women  **OR**  On one of the following drug treatments for reduced HDL cholesterol, based on participant self-reported medication data:   - Naftidrofuryl Oxalate - Niacin - Pentoxifylline - Simvastatin - Atorvastatin - Rosuvastatin - Pravastatin - Fluvastatin - Fenofibrate - Bezafibrate - Ciprofibrate - Gemfibrozil - Acipimox | 3076, 20003 |
| Hyperglycaemia | Fasting glucose ⩾100 mg/dL  **OR**  On one of the following drug treatments for elevated glucose, based on participant self-reported medication data:   - Metformin - Insulin - Gliclazide - Pioglitazone - Rosiglitazone - Glimepiride - Metformin \|Rosiglitazone - Glyburide - Glipizide - Repaglinide - Tolbutamide - Acarbose - Nateglinide | 30740, 20003 |
| Metabolic syndrome | Case status for at least three of the five metabolic outcomes defined above:   1. High waist circumference 2. Hypertension 3. Hypertriglyceridemia 4. Low HDL cholesterol 5. Hyperglycemia | All field codes listed in rows above |

**Supplementary Table 4:** Full associations between each candidate SNP and each of the 6 phenotypic definitions for depression. Regression models were adjusted for age, age^2^, sex, genotyping batch, testing centre, and the first six European ancestry principal components. Unadjusted p values and q-values (calculated by applying false discovery rate correction across phenotype definitions) are presented.

| **SNP** | **A1/A2** | **Broad** | | | **ICD10-coded** | | | **Lifetime** | | | **CIDI** | | | **PHQ-9 definition** | | | **PHQ-9 cutoff** | | |
| --- | --- | --- | --- | --- | --- | --- | --- | --- | --- | --- | --- | --- | --- | --- | --- | --- | --- | --- | --- |
|  |  | **B (Std. Error)** | ***P*** | **q-value** | **B (Std. Error)** | ***P*** | **q-value** | **B (Std. Error)** | ***P*** | **q-value** | **B (Std. Error)** | ***P*** | **q-value** | **B (Std. Error)** | ***P*** | **q-value** | **B (Std. Error)** | ***P*** | **q-value** |
| rs1982632 | A/G | 0.00131 (0.00639) | 0.837 | 0.954 | -0.00079 (0.0137) | 0.954 | 0.954 | 0.0017 (0.0143) | 0.905 | 0.954 | -0.0142 (0.0122) | 0.247 | 0.74 | -0.0185 (0.0257) | 0.473 | 0.946 | -0.0407 (0.0254) | 0.109 | 0.652 |
| rs78161395 | T/G | -0.0139 (0.00684) | 0.0428 | 0.257 | -0.003 (0.0147) | 0.838 | 0.919 | -0.0112 (0.0152) | 0.463 | 0.919 | -0.00133 (0.013) | 0.919 | 0.919 | 0.0105 (0.0272) | 0.698 | 0.919 | 0.0163 (0.0266) | 0.539 | 0.919 |
| rs74400983 | T/C | 0.0248 (0.0109) | 0.0232 | 0.139 | 0.0206 (0.0232) | 0.376 | 0.654 | 0.0369 (0.0242) | 0.127 | 0.381 | -0.0127 (0.0209) | 0.545 | 0.654 | -0.0323 (0.044) | 0.462 | 0.654 | -0.0136 (0.043) | 0.752 | 0.752 |
| rs6511905 | G/C | -0.00102 (0.00587) | 0.862 | 0.989 | 0.00367 (0.0126) | 0.77 | 0.989 | 0.000182 (0.0131) | 0.989 | 0.989 | -0.00582 (0.0112) | 0.604 | 0.989 | 0.00214 (0.0234) | 0.927 | 0.989 | 0.00387 (0.0229) | 0.866 | 0.989 |
| rs9292519 | A/G | -0.00883 (0.00508) | 0.0822 | 0.247 | -0.0105 (0.0109) | 0.335 | 0.67 | -0.00722 (0.0113) | 0.524 | 0.787 | -0.0182 (0.00973) | 0.0611 | 0.247 | -0.00631 (0.0203) | 0.756 | 0.865 | 0.00338 (0.0199) | 0.865 | 0.865 |
| rs171631 | A/C | 0.00469 (0.0106) | 0.658 | 0.789 | -0.0192 (0.0229) | 0.402 | 0.604 | 0.00343 (0.0236) | 0.885 | 0.885 | -0.0384 (0.0203) | 0.0582 | 0.349 | -0.0418 (0.0429) | 0.33 | 0.604 | -0.04 (0.0421) | 0.342 | 0.604 |
| rs42868 | G/C | 0.00317 (0.00684) | 0.643 | 0.749 | -0.00515 (0.0147) | 0.725 | 0.749 | -0.0163 (0.0153) | 0.286 | 0.749 | -0.0167 (0.0131) | 0.203 | 0.749 | 0.00869 (0.0272) | 0.749 | 0.749 | 0.0225 (0.0266) | 0.398 | 0.749 |
| rs7702361 | A/C | 0.00836 (0.00509) | 0.1 | 0.217 | 0.00462 (0.0109) | 0.672 | 0.806 | 0.0183 (0.0114) | 0.108 | 0.217 | 0.0191 (0.00974) | 0.0499 | 0.217 | 0.00065 (0.0203) | 0.974 | 0.974 | -0.0219 (0.02) | 0.274 | 0.41 |
| rs11264422 | T/A | 0.00591 (0.00528) | 0.263 | 0.621 | 0.0114 (0.0113) | 0.311 | 0.621 | 0.0155 (0.0118) | 0.187 | 0.621 | -0.00405 (0.0101) | 0.688 | 0.825 | -0.00386 (0.021) | 0.854 | 0.854 | -0.00854 (0.0207) | 0.679 | 0.825 |
| rs62351166 | A/C | 0.0135 (0.00656) | 0.0388 | 0.233 | 0.0148 (0.014) | 0.29 | 0.435 | 0.0156 (0.0147) | 0.289 | 0.435 | 0.000899 (0.0126) | 0.943 | 0.943 | -0.0299 (0.0267) | 0.262 | 0.435 | 0.00487 (0.0259) | 0.851 | 0.943 |
| rs7695640 | G/A | 0.00748 (0.00719) | 0.298 | 0.894 | 0.000788 (0.0154) | 0.959 | 0.959 | 0.00268 (0.0161) | 0.868 | 0.959 | 0.0193 (0.0138) | 0.16 | 0.894 | -0.0125 (0.029) | 0.665 | 0.959 | -0.0177 (0.0285) | 0.535 | 0.959 |
| rs11100192 | G/A | 0.0225 (0.0225) | 0.318 | 0.747 | 0.0352 (0.0475) | 0.458 | 0.747 | 0.0457 (0.05) | 0.361 | 0.747 | -0.0147 (0.0428) | 0.731 | 0.747 | 0.0285 (0.0885) | 0.747 | 0.747 | -0.032 (0.089) | 0.719 | 0.747 |
| rs72703633 | C/T | 0.0187 (0.0234) | 0.423 | 0.896 | 0.0259 (0.0497) | 0.603 | 0.896 | 0.0485 (0.0511) | 0.343 | 0.896 | 0.00877 (0.0443) | 0.843 | 0.896 | 0.012 (0.0916) | 0.896 | 0.896 | 0.0518 (0.0889) | 0.56 | 0.896 |
| rs11793069 | G/A | -0.0027 (0.00503) | 0.592 | 0.887 | -0.018 (0.0108) | 0.0946 | 0.568 | -0.00144 (0.0112) | 0.898 | 0.898 | 0.00399 (0.00959) | 0.677 | 0.887 | 0.00905 (0.02) | 0.651 | 0.887 | -0.00654 (0.0197) | 0.739 | 0.887 |
| rs72499174 | C/G | -0.0054 (0.00589) | 0.36 | 0.67 | 0.0213 (0.0126) | 0.0902 | 0.541 | 0.0105 (0.0131) | 0.421 | 0.67 | -0.00855 (0.0112) | 0.447 | 0.67 | 0.00444 (0.0234) | 0.849 | 0.849 | 0.00591 (0.023) | 0.797 | 0.849 |

**Supplementary Table 5:** Full associations between each candidate SNP and each of the 3 phenotypic definitions for atypical depression. Regression models were adjusted for age, age^2^, sex, genotyping batch, testing centre, and the first six European ancestry principal components. Unadjusted p values and q-values (calculated by applying false discovery rate correction across phenotype definitions) are presented.

| **SNP** | **A1/A2** | **CIDI** | | | **PHQ-9 definition** | | | **PHQ-9 cutoff** | | |
| --- | --- | --- | --- | --- | --- | --- | --- | --- | --- | --- |
|  |  | **B (Std. Error)** | ***P*** | **q-value** | **B (Std. Error)** | ***P*** | **q-value** | **B (Std. Error)** | ***P*** | **q-value** |
| rs1982632 | A/G | 0.0266 (0.0453) | 0.557 | 0.836 | 0.0651 (0.0875) | 0.457 | 0.836 | -0.0157 (0.0782) | 0.841 | 0.841 |
| rs78161395 | T/G | 0.0389 (0.0483) | 0.42 | 0.63 | 0.0817 (0.0924) | 0.376 | 0.63 | -0.0338 (0.084) | 0.687 | 0.687 |
| rs74400983 | T/C | -0.0239 (0.0785) | 0.761 | 0.761 | -0.304 (0.174) | 0.0804 | 0.241 | -0.128 (0.141) | 0.364 | 0.546 |
| rs6511905 | G/C | 0.0164 (0.0417) | 0.695 | 0.831 | 0.0173 (0.081) | 0.831 | 0.831 | -0.0245 (0.0716) | 0.732 | 0.831 |
| rs9292519 | A/G | -0.0295 (0.0365) | 0.418 | 0.82 | 0.0161 (0.0707) | 0.820 | 0.820 | 0.0278 (0.0616) | 0.652 | 0.82 |
| rs171631 | A/C | -0.00155 (0.0754) | 0.984 | 0.984 | -0.153 (0.157) | 0.329 | 0.984 | -0.0248 (0.13) | 0.848 | 0.984 |
| rs42868 | G/C | -0.0298 (0.0493) | 0.546 | 0.546 | -0.154 (0.1) | 0.125 | 0.188 | -0.193 (0.089) | 0.0297 | 0.089 |
| rs7702361 | A/C | 0.0304 (0.0365) | 0.404 | 0.770 | 0.0208 (0.0709) | 0.770 | 0.770 | 0.0381 (0.0622) | 0.540 | 0.77 |
| rs11264422 | T/A | -0.0348 (0.0378) | 0.357 | 0.536 | -0.0216 (0.0734) | 0.769 | 0.769 | 0.0669 (0.0634) | 0.291 | 0.536 |
| rs62351166 | A/C | 0.0525 (0.0466) | 0.260 | 0.756 | -0.0088 (0.0921) | 0.924 | 0.924 | 0.0526 (0.0788) | 0.504 | 0.756 |
| rs7695640 | G/A | 0.0632 (0.0508) | 0.214 | 0.568 | -0.072 (0.103) | 0.486 | 0.568 | 0.0494 (0.0865) | 0.568 | 0.568 |
| rs11100192 | G/A | 0.0344 (0.156) | 0.825 | 0.854 | -0.0592 (0.321) | 0.854 | 0.854 | -0.0764 (0.282) | 0.786 | 0.854 |
| rs72703633 | C/T | -0.185 (0.183) | 0.311 | 0.484 | 0.2 (0.293) | 0.494 | 0.494 | 0.252 (0.254) | 0.323 | 0.484 |
| rs11793069 | G/A | 0.0216 (0.0358) | 0.547 | 0.937 | -0.0223 (0.07) | 0.75 | 0.937 | -0.00479 (0.0608) | 0.937 | 0.937 |
| rs72499174 | C/G | 0.0211 (0.0417) | 0.612 | 0.894 | -0.0975 (0.0839) | 0.245 | 0.736 | -0.0095 (0.0715) | 0.894 | 0.894 |

**Supplementary Table 6:** Full associations between each candidate SNP and each of the 3 phenotypic definitions for anxiety disorders. Regression models were adjusted for age, age^2^, sex, genotyping batch, testing centre, and the first six European ancestry principal components. Unadjusted p values and q-values (calculated by applying false discovery rate correction across phenotype definitions) are presented.

| **SNP** | **A1/A2** | **ICD10-coded** | | | **Probable** | | | **GAD-7 cutoff** | | |
| --- | --- | --- | --- | --- | --- | --- | --- | --- | --- | --- |
|  |  | **B (Std. Error)** | ***P*** | **q-value** | **B (Std. Error)** | ***P*** | **q-value** | **B (Std. Error)** | ***P*** | **q-value** |
| rs1982632 | A/G | 0.0110 (0.0148) | 0.455 | 0.455 | 0.0141 (0.0143) | 0.322 | 0.455 | -0.0254 (0.0282) | 0.367 | 0.455 |
| rs78161395 | T/G | -0.000691 (0.0158) | 0.965 | 0.965 | -0.00866 (0.0153) | 0.571 | 0.942 | -0.0146 (0.0300) | 0.628 | 0.942 |
| rs74400983 | T/C | -0.00715 (0.0254) | 0.778 | 0.778 | -0.0193 (0.0245) | 0.432 | 0.686 | -0.036 (0.0485) | 0.457 | 0.686 |
| rs6511905 | G/C | -0.00906 (0.0136) | 0.506 | 0.506 | -0.0136 (0.0131) | 0.300 | 0.450 | -0.0295 (0.0259) | 0.254 | 0.450 |
| rs9292519 | A/G | -0.0137 (0.0118) | 0.245 | 0.736 | -0.00178 (0.0114) | 0.876 | 0.904 | -0.00268 (0.0223) | 0.904 | 0.904 |
| rs171631 | A/C | -0.0248 (0.0248) | 0.356 | 0.534 | 0.0216 (0.0234) | 0.356 | 0.534 | -0.000426 (0.0462) | 0.993 | 0.993 |
| rs42868 | G/C | 0.00378 (0.0158) | 0.811 | 0.811 | -0.0104 (0.0153) | 0.496 | 0.743 | -0.0328 (0.0303) | 0.279 | 0.743 |
| rs7702361 | A/C | 0.00478 (0.0118) | 0.685 | 0.685 | 0.00640 (0.0114) | 0.574 | 0.685 | 0.0241 (0.0223) | 0.280 | 0.685 |
| rs11264422 | T/A | 0.0311 (0.0122) | 0.0106 | 0.016 | -0.0348 (0.0118) | 0.00318 | 0.010 | -0.0101 (0.0231) | 0.661 | 0.661 |
| rs62351166 | A/C | 0.00868 (0.0152) | 0.568 | 0.568 | 0.0253 (0.0147) | 0.0856 | 0.257 | -0.0229 (0.0292) | 0.433 | 0.568 |
| rs7695640 | G/A | 0.00132 (0.0166) | 0.937 | 0.937 | 0.00372 (0.0161) | 0.817 | 0.937 | -0.00431 (0.0317) | 0.892 | 0.937 |
| rs11100192 | G/A | -0.0516 (0.0534) | 0.334 | 0.906 | 0.00593 (0.0500) | 0.906 | 0.906 | 0.0170 (0.0970) | 0.861 | 0.906 |
| rs72703633 | C/T | 0.0783 (0.0526) | 0.137 | 0.376 | 0.0105 (0.0519) | 0.840 | 0.840 | 0.111 (0.0966) | 0.251 | 0.376 |
| rs11793069 | G/A | 0.00324 (0.0117) | 0.781 | 0.781 | -0.0180 (0.0112) | 0.108 | 0.325 | 0.0215 (0.0220) | 0.329 | 0.493 |
| rs72499174 | C/G | 0.00871 (0.0136) | 0.523 | 0.821 | 0.00734 (0.0131) | 0.575 | 0.821 | 0.00581 (0.0257) | 0.821 | 0.821 |

| **SNP** | **A1/A2** | **Metabolic syndrome** | | | **Hypertension** | | | **Hypertriglyceridaemia** | | | **Low HDL Cholesterol**  **(Dyslipidaemia)** | | | **Hyperglycaemia** | | |
| --- | --- | --- | --- | --- | --- | --- | --- | --- | --- | --- | --- | --- | --- | --- | --- | --- |
|  |  | **B (Std. Error)** | ***P*** | **q-value** | **B (Std. Error)** | ***P*** | **q-value** | **B (Std. Error)** | ***P*** | **q-value** | **B (Std. Error)** | ***P*** | **q-value** | **B (Std. Error)** | ***P*** | **q-value** |
| rs1982632 | A/G | 0.00711 (0.00722) | 0.325 | 0.568 | 0.00801 (0.00739) | 0.278 | 0.568 | 0.00597 (0.00668) | 0.372 | 0.568 | -0.00599 (0.008) | 0.454 | 0.568 | 0.00571 (0.012) | 0.633 | 0.633 |
| rs78161395 | T/G | -0.00229 (0.00771) | 0.766 | 0.8 | -0.00295 (0.00786) | 0.708 | 0.8 | 0.00204 (0.00712) | 0.775 | 0.8 | 0.00216 (0.00853) | 0.8 | 0.8 | -0.015 (0.0128) | 0.244 | 0.800 |
| rs74400983 | T/C | -0.000772 (0.0123) | 0.95 | 0.995 | -0.000082 (0.0126) | 0.995 | 0.995 | -0.00849 (0.0114) | 0.457 | 0.867 | 0.00876 (0.0136) | 0.52 | 0.867 | -0.0198 (0.0206) | 0.336 | 0.867 |
| rs6511905 | G/C | -0.00326 (0.00662) | 0.623 | 0.779 | -0.00189 (0.00675) | 0.779 | 0.779 | 0.00305 (0.00612) | 0.618 | 0.779 | 0.0048 (0.00732) | 0.512 | 0.779 | -0.0157 (0.011) | 0.154 | 0.768 |
| rs9292519 | A/G | -0.00726 (0.00573) | 0.205 | 0.721 | -0.000269 (0.00585) | 0.963 | 0.963 | 0.00053 (0.0053) | 0.92 | 0.963 | -0.0061 (0.00635) | 0.337 | 0.721 | -0.00747 (0.00952) | 0.433 | 0.721 |
| rs171631 | A/C | -0.00176 (0.0119) | 0.882 | 0.972 | -0.00042 (0.0122) | 0.972 | 0.972 | -0.0105 (0.0111) | 0.343 | 0.857 | -0.0244 (0.0133) | 0.0663 | 0.331 | -0.00142 (0.0198) | 0.943 | 0.972 |
| rs42868 | G/C | -0.00507 (0.00773) | 0.512 | 0.512 | 0.013 (0.0079) | 0.0988 | 0.165 | -0.0144 (0.00715) | 0.0445 | 0.111 | -0.0188 (0.00859) | 0.0289 | 0.111 | -0.00875 (0.0128) | 0.496 | 0.512 |
| rs7702361 | A/C | 0.0078 (0.00575) | 0.175 | 0.437 | -0.00606 (0.00587) | 0.302 | 0.503 | 0.002 (0.00532) | 0.707 | 0.707 | 0.00937 (0.00636) | 0.141 | 0.437 | 0.00759 (0.00953) | 0.425 | 0.532 |
| rs11264422 | T/A | -0.00233 (0.00595) | 0.696 | 0.87 | -0.00598 (0.00608) | 0.325 | 0.542 | 0.00951 (0.00551) | 0.0846 | 0.251 | 0.0108 (0.00659) | 0.101 | 0.251 | -0.00075 (0.00988) | 0.939 | 0.939 |
| rs62351166 | A/C | 0.00453 (0.00742) | 0.542 | 0.542 | 0.0055 (0.00757) | 0.468 | 0.542 | 0.00934 (0.00685) | 0.173 | 0.542 | -0.00754 (0.00821) | 0.358 | 0.542 | -0.0129 (0.0123) | 0.297 | 0.542 |
| rs7695640 | G/A | -0.00726 (0.00812) | 0.371 | 0.371 | -0.0102 (0.00829) | 0.219 | 0.293 | -0.0125 (0.00752) | 0.0953 | 0.293 | -0.0107 (0.00902) | 0.235 | 0.293 | -0.0163 (0.0135) | 0.227 | 0.293 |
| rs11100192 | G/A | 0.00814 (0.0256) | 0.75 | 0.75 | -0.0266 (0.0259) | 0.304 | 0.539 | -0.0255 (0.0237) | 0.281 | 0.539 | 0.0227 (0.0281) | 0.418 | 0.539 | 0.033 (0.0419) | 0.431 | 0.539 |
| rs72703633 | C/T | 0.00986 (0.0264) | 0.709 | 0.824 | 0.0397 (0.027) | 0.141 | 0.706 | -0.0143 (0.0247) | 0.562 | 0.824 | -0.0119 (0.0295) | 0.686 | 0.824 | -0.00983 (0.0441) | 0.824 | 0.824 |
| rs11793069 | G/A | 0.00958 (0.00567) | 0.0913 | 0.228 | 0.0122 (0.0058) | 0.0353 | 0.176 | 0.00686 (0.00525) | 0.192 | 0.319 | 0.00298 (0.00628) | 0.635 | 0.635 | -0.00739 (0.00941) | 0.432 | 0.540 |
| rs72499174 | C/G | -0.00974 (0.00665) | 0.143 | 0.238 | -0.0182 (0.00677) | 0.00709 | 0.0355 | -0.0135 (0.00616) | 0.0282 | 0.0704 | -0.00165 (0.00736) | 0.823 | 0.823 | 0.0052 (0.011) | 0.637 | 0.796 |

**Supplementary Table 7:** Full associations between each candidate SNP and metabolic syndrome, as well as 4 sub-outcomes that comprise metabolic syndrome. Regression models were adjusted for age, age^2^, sex, genotyping batch, testing centre, and the first six European ancestry principal components. Unadjusted p values and q-values (calculated by applying false discovery rate correction across phenotype definitions) are presented.

**Supplementary Table 8:** Results of a multivariate regression model with all candidate SNPs at a particular gene as simultaneous explanatory variables and several depression and atypical depression phenotype outcomes, adjusted for age, age^2^, sex, genotyping batch, testing centre, and the first six European ancestry principal components. Unadjusted p values and q-values (calculated by applying false discovery rate correction across phenotype definitions) are presented.

| **Phenotype** | **Gene** | **Chi-square** | ***P*** | **q-value** |
| --- | --- | --- | --- | --- |
| Broad depression | RLN3 | 10.00 | 0.0404 | 0.242 |
|  | RXFP3 | 5.31 | 0.257 | 0.745 |
|  | RXFP1 | 7.14 | 0.0676 | 0.406 |
|  | RLN2 | 2.64 | 0.451 | 0.934 |
| ICD10-coded depression | RLN3 | 0.62 | 0.961 | 0.961 |
|  | RXFP3 | 1.95 | 0.745 | 0.745 |
|  | RXFP1 | 1.36 | 0.715 | 0.859 |
|  | RLN2 | 4.23 | 0.238 | 0.934 |
| Lifetime depression | RLN3 | 3.26 | 0.516 | 0.927 |
|  | RXFP3 | 3.08 | 0.544 | 0.745 |
|  | RXFP1 | 2.50 | 0.476 | 0.859 |
|  | RLN2 | 2.02 | 0.569 | 0.934 |
| CIDI depression | RLN3 | 2.65 | 0.618 | 0.927 |
|  | RXFP3 | 5.90 | 0.206 | 0.745 |
|  | RXFP1 | 2.97 | 0.397 | 0.859 |
|  | RLN2 | 0.71 | 0.871 | 0.934 |
| PHQ-9 definition depression | RLN3 | 1.64 | 0.801 | 0.961 |
|  | RXFP3 | 2.40 | 0.663 | 0.745 |
|  | RXFP1 | 1.71 | 0.636 | 0.859 |
|  | RLN2 | 0.43 | 0.934 | 0.934 |
| PHQ-9 cutoff depression | RLN3 | 2.99 | 0.559 | 0.927 |
|  | RXFP3 | 4.06 | 0.398 | 0.745 |
|  | RXFP1 | 0.50 | 0.92 | 0.920 |
|  | RLN2 | 0.61 | 0.894 | 0.934 |
| CIDI atypical depression | RLN3 | 1.78 | 0.777 | 0.777 |
|  | RXFP3 | 4.69 | 0.321 | 0.481 |
|  | RXFP1 | 3.10 | 0.377 | 0.871 |
|  | RLN2 | 2.11 | 0.55 | 0.796 |
| PHQ-9 definition atypical depression | RLN3 | 6.38 | 0.173 | 0.516 |
|  | RXFP3 | 2.88 | 0.578 | 0.578 |
|  | RXFP1 | 0.71 | 0.871 | 0.871 |
|  | RLN2 | 2.79 | 0.426 | 0.796 |
| PHQ-9 cutoff atypical depression | RLN3 | 4.49 | 0.344 | 0.516 |
|  | RXFP3 | 5.75 | 0.219 | 0.481 |
|  | RXFP1 | 1.27 | 0.737 | 0.871 |
|  | RLN2 | 1.02 | 0.796 | 0.796 |

**Supplementary Table 9:** Results of a multivariate regression model with all candidate SNPs at a particular gene as simultaneous explanatory variables and several anxiety phenotype outcomes, adjusted for age, age^2^, sex, genotyping batch, testing centre, and the first six European ancestry principal components. Unadjusted p values and q-values (calculated by applying false discovery rate correction across phenotype definitions) are presented.

| **Phenotype** | **Gene** | **Chi-square** | ***P*** | **q-value** |
| --- | --- | --- | --- | --- |
| ICD10-coded anxiety | RLN3 | 2.54 | 0.638 | 0.676 |
|  | RXFP3 | 2.91 | 0.573 | 0.676 |
|  | RXFP1 | 1.53 | 0.676 | 0.676 |
|  | RLN2 | 2.08 | 0.556 | 0.794 |
| Lifetime disorder anxiety | RLN3 | 1.68 | 0.794 | 0.794 |
|  | RXFP3 | 8.18 | 0.0853 | 0.256 |
|  | RXFP1 | 2.97 | 0.396 | 0.566 |
|  | RLN2 | 2.69 | 0.442 | 0.566 |
| GAD-7 cutoff anxiety | RLN3 | 2.95 | 0.566 | 0.566 |
|  | RXFP3 | 2.41 | 0.661 | 0.856 |
|  | RXFP1 | 0.77 | 0.856 | 0.856 |
|  | RLN2 | 2.27 | 0.518 | 0.856 |

**Supplementary Table 10:** Results of a multivariate regression model with all candidate SNPs at a particular gene as simultaneous explanatory variables and metabolic syndrome, as well as the sub-outcomes comprising the disorder, adjusted for age, age^2^, sex, genotyping batch, testing centre, and the first six European ancestry principal components.

| **Phenotype** | **Gene** | **Chi-square** | ***P*** | **q-value** |
| --- | --- | --- | --- | --- |
| Metabolic syndrome | RLN3 | 1.34 | 0.854 | 0.894 |
|  | RXFP3 | 2.68 | 0.613 | 0.766 |
|  | RXFP1 | 1.12 | 0.773 | 0.773 |
|  | RLN2 | 3.41 | 0.333 | 0.555 |
| Hypertension | RLN3 | 1.37 | 0.85 | 0.894 |
|  | RXFP3 | 7.60 | 0.107 | 0.521 |
|  | RXFP1 | 2.38 | 0.497 | 0.622 |
|  | RLN2 | 11.20 | 0.0109 | 0.054 |
| Hypertriglyceridaemia | RLN3 | 6.89 | 0.142 | 0.710 |
|  | RXFP3 | 4.76 | 0.313 | 0.521 |
|  | RXFP1 | 5.63 | 0.131 | 0.589 |
|  | RLN2 | 5.62 | 0.132 | 0.330 |
| Low HDL cholesterol  (Dyslipidaemia) | RLN3 | 1.10 | 0.894 | 0.894 |
|  | RXFP3 | 4.86 | 0.302 | 0.521 |
|  | RXFP1 | 3.26 | 0.353 | 0.589 |
|  | RLN2 | 0.53 | 0.912 | 0.912 |
| Hyperglycaemia | RLN3 | 4.30 | 0.367 | 0.894 |
|  | RXFP3 | 1.06 | 0.901 | 0.901 |
|  | RXFP1 | 3.60 | 0.308 | 0.589 |
|  | RLN2 | 0.77 | 0.856 | 0.912 |
